# COGNITION AND INFLUENCING FACTORS OF HOSPICE CARE AMONG NURSING UNDERGRADUATES

**DOI:** 10.1101/2022.07.27.22278100

**Authors:** Rui-ting Zhang

## Abstract

**Objective:** To investigate the current cognition of hospice care among nursing undergraduates, and analyze the influencing factors affecting their cognition, so as to provide suggestions for improving the cognition of hospice care among nursing undergraduates.

**Methods:** 290 nursing undergraduates were selected for an online questionnaire survey. The questionnaire includes general information, hospice care knowledge, and hospice care attitude.

**Results:** The mean score of the hospice care knowledge questionnaire is 9.68±4.73, and the mean score of the attitude questionnaire is 62.59±7.29. The influencing factors of hospice care cognition of nursing undergraduates are gender, age, religious belief, the only child, internship experience, grade, source of death experience, death talk at home, and access to hospice care knowledge.

**Conclusion:** Nursing undergraduates’ lack of knowledge of hospice care, and hospice care attitude is more positive, suggesting that nursing undergraduates should strengthen hospice care education according to the influencing factors, call for government support, in order to further improve nursing undergraduates’ hospice care cognition status.

---

According to Health China 2030, The theme of health in the next decade is to cover the whole life cycle, identify a number of priority areas according to the main health problems and main influencing factors in different stages of life, strengthen intervention, realize the whole process of health services and health security from the fetus to the end of life, and comprehensively safeguard people’s health^[1]^.Like the 10 months of maternal womb care, the end of life also needs 10 months of social care, that is, the whole society to create an atmosphere similar to the care of the fetus, so that people in the final phase of warmth and love^[2]^That’s hospice care. Hospice care can alleviate the physical pain, especially the mental pain, improve the quality of death and achieve optimal death.

Among medical staff, nursing staff have more time to contact with terminal patients and are the main force of nursing staff. Previous studies have shown that the cognitive level of hospice care of nursing staff directly affects the quality of hospice care^[3-6]^.Undergraduate nursing students as engaged in hospice nursing with higher education, their hospice care cognition can not be ignored, undergraduate nursing students to hospice care cognition and attitude will directly affect their future hospice care service behavior^[7]^.Previous studies have shown that nursing undergraduates’ cognition of hospice care needs to be strengthened^[8-13]^Therefore, in order to effectively improve the cognition of hospice care for nursing undergraduates, it is necessary to understand the influencing factors of hospice care cognition for nursing undergraduates, so as to further adopt reasonable methods to improve cognition for the influencing factors.

## 1 Objects and Methods

### 1.1 Object

290 nursing undergraduates were selected as research subjects with 5 times the number of questionnaire items and 20% shedding rate estimated. Inclusion criteria: ① Undergraduate nursing students, ② Informed consent and voluntary participation in the study, ③ Can correctly understand the meaning of each item in the questionnaire. Exclusion criteria: incomplete questionnaire filling.

### 1.2 Methods

#### 1.2.1 Survey Tools

The questionnaire includes three parts: general information, hospice knowledge, and hospice attitude. The general information questionnaire was designed by ourselves after the literature review, including 10 items such as gender, grade, age, religious belief, and internship experience.

The end-of-life care knowledge questionnaire was quoted from the end-of-life care knowledge questionnaire designed by the Meng Na team in 2017, which was modified by the Meng Na team by combining the ROSS palliative care knowledge questionnaire with the PCQN Chinese scale translated by Zou Min in 2007^[14],^ a total of 18 questions with 4 dimensions, 1 point is scored for each question, 0 points are scored for each question wrong or uncertain, and the final score is the sum of the scores of 18 questions. The higher the score is, the better the knowledge of hospice care is mastered; otherwise, the worse it is.

The hospice Attitude questionnaire was quoted from the hospice Attitude Questionnaire designed by the Meng Na team in 2017, which was modified by the Meng Na team by combining Naomi R. Adelman’s hospice Primary Caregiver questionnaire, Huang Tian zhong’s Death Attitude Questionnaire, and Zou Min’s attitude assessment questionnaire in 2003^[14]^, including 19 topics and 3 dimensions. It includes 10 positive questions and 9 negative questions. The score of 10 positive questions is 5 points, and the score of the remaining questions is successively decreased. The score of 9 negative questions is 1 point, the score of the remaining questions is successively increased, and the final score is the sum of the scores of 19 questions.

#### 1.2.2 Data Collection Method

The questionnaire can be sent and returned online only once for the same mobile phone and IP address.

#### 1.2.3 Statistical Analysis Methods

SPSS20.0 was used for data analysis. Mean ± standard deviation 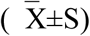 was used to describe the measurement data, frequency and constituent ratio were used to describe the counting data, one-way an OVA was used to analyze the influencing factors of cognition, multiple linear regression was used to analyze the independent influencing factors, P<0.05 was considered statistically significant. ^−^

## 2 Results

### 2.1. General Data

In the 290 valid questionnaires collected, about one-third of the respondents are senior students, and most of them are female students. Most of them have internship experience, most of them have no religious belief, and more than half of them are only children with an average age of 20(Table 1).

**Table 1.** Demographic data

|  |  | Frequency | Proportion |
| --- | --- | --- | --- |
| Gender | Male | 109 | 37.59% |
|  | Female | 181 | 62.41% |
| Religion | Yes | 34 | 11.72% |
|  | No | 256 | 88.28% |
| The only child in the family | Yes | 171 | 58.97% |
|  | No | 119 | 41.03% |
| Internship Experience | Yes | 188 | 64.83% |
|  | No | 102 | 35.17% |
| Grade | Freshman | 45 | 13.11% |
|  | Sophomore | 39 | 13.44% |
|  | Junior | 76 | 26.21% |
|  | Senior | 108 | 47.24% |

#### 2.1.1 Sources of Nursing Undergraduate Death Experience

42.07% of nursing undergraduates’ death experiences came from grandparents, 24.48% from parents, and the rest from strangers etc (Figure 1).

**Figure 1.**
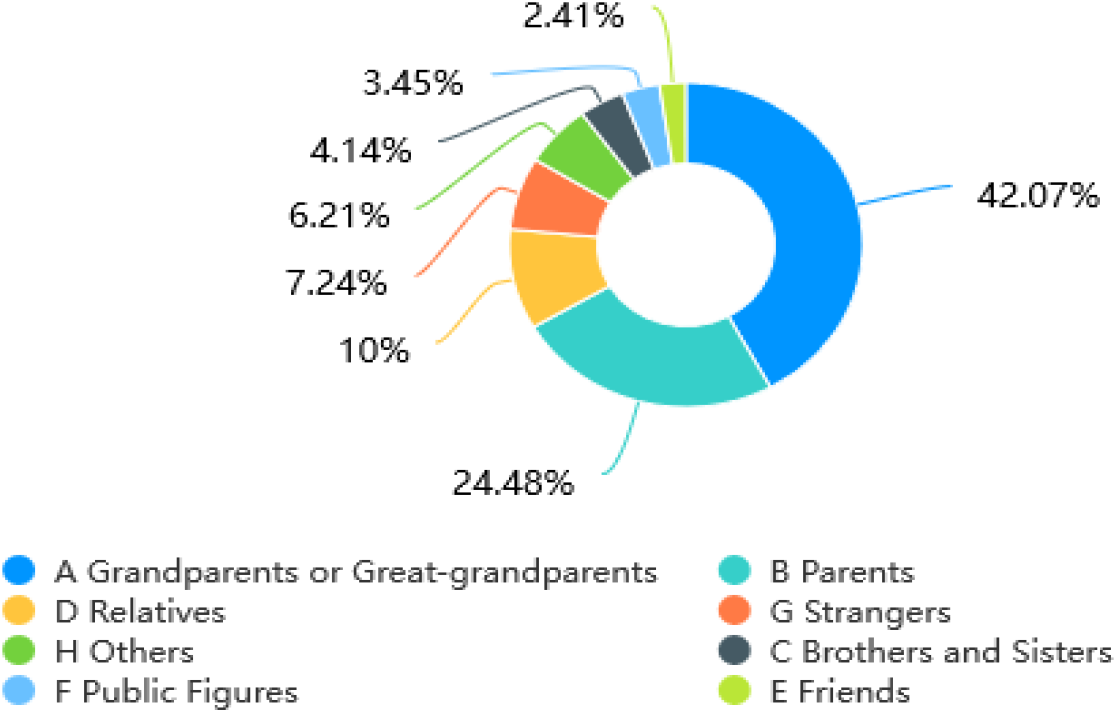

#### 2.1.2 Discussion on Death Situation of Nursing Undergraduates at Home

26.5% of surveyed families felt uncomfortable discussing death, 25.8% of surveyed families openly discussed death, 21% of families avoided talking about death, and 16.5% of families only talked about this topic when necessary and avoided children(Figure 2).

**Figure 2.**
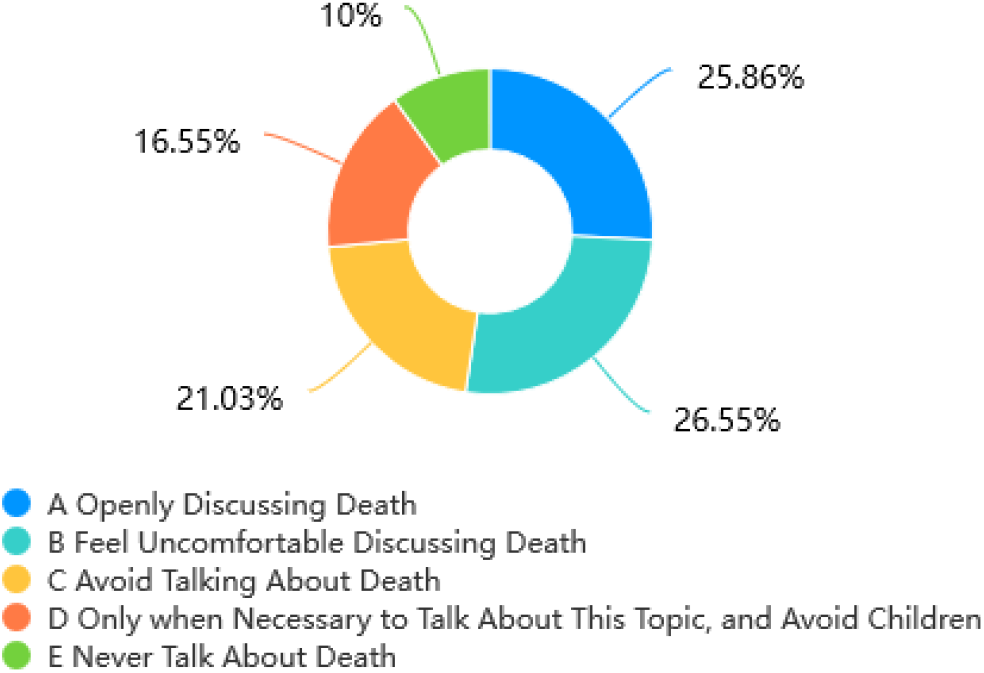

#### 2.1.3 Sources of Hospice Care Knowledge for Nursing Undergraduates

37.93% of nursing undergraduates’ hospice care knowledge came from classroom, 19.31% from media, and the rest from professional books, lectures and social activities(Figure 3).

**Figure 3.**
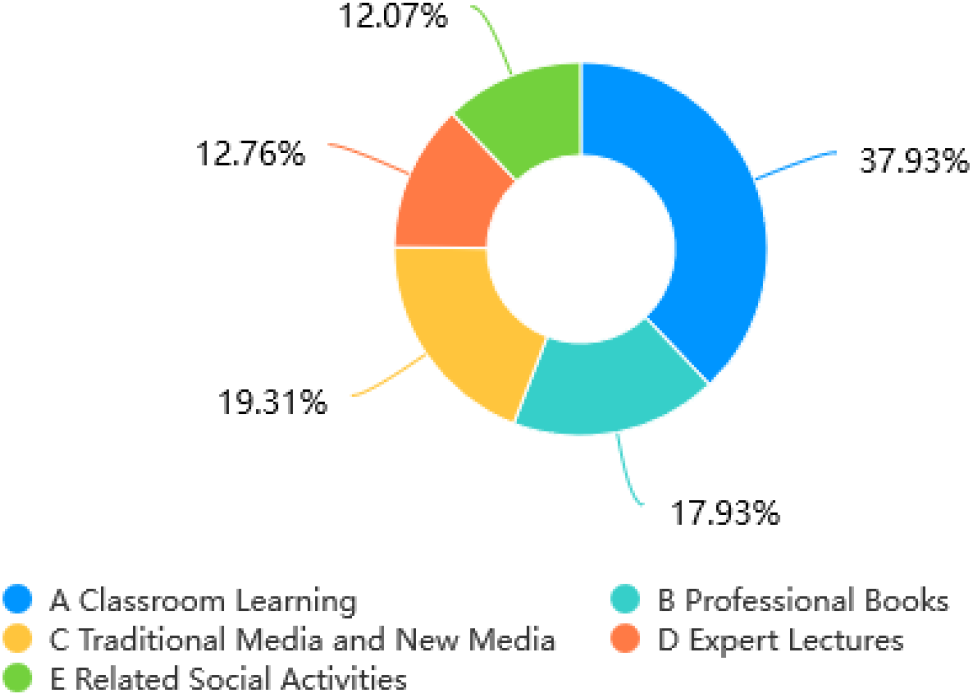

#### 2.1.4 Channels for Nursing Undergraduates to Acquire Hospice Care Knowledge

56.5% of nursing undergraduates expected to acquire the knowledge of end-of-life care from classroom learning, 55.9% from professional books, 51.4% from media, 48.9% from lectures, 41.7% of nursing undergraduates hoped to gain hospice care knowledge from social activities(Figure 4).

**Figure 4.**
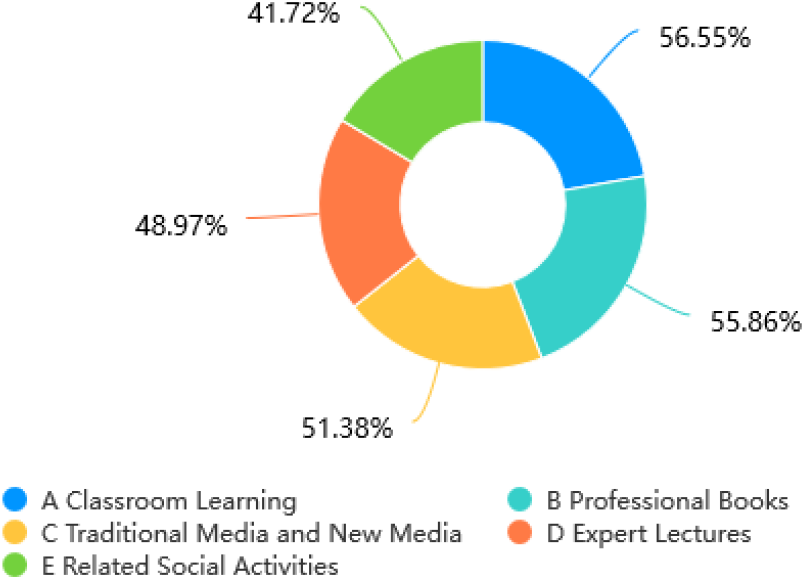

### 2.2 Analysis of Hospice Knowledge Questionnaire

#### 2.2.1 Score of Hospice Knowledge Questionnaire for Nursing Undergraduates

There are 18 questions in this questionnaire, 1 point for each question, and the total score is 18 points. In all valid questionnaires, the highest score was 18, the lowest score was 0, the average score was 9.6793±4.7340, and the average accuracy rate was 53.77%(Table 2).

**Table 2.** Score of Hospice Knowledge Questionnaire

| Dimension | Mean Score | Standard Deviation |
| --- | --- | --- |
| Basic Knowledge | 1.6586 | 0.8251 |
| Philosophy and Principles | 1.7166 | 0.8116 |
| Pain Control | 1.7474 | 0.8114 |
| Social ,Psychological and Spiritual Support | 1.6448 | 0.8218 |

#### 2.2.2 Single Factor Analysis of Influencing Factors of Hospice Care Knowledge of Nursing Undergraduates

The one-way ANOVA was used to analyze the scores of four dimensions of the hospice care knowledge questionnaire: basic knowledge, philosophy and principles, pain control, social, psychological and spiritual support.

Dimension of basic knowledge: The scores of whether the child was the only child, different grades, different sources of death experience, and whether or not the student had internship were statistically significant (P < 0.05). The scores of hospice care knowledge of nursing undergraduates who were the only child, the freshman, and the sources of death experience from parents and internship were higher.

Philosophy and principles dimension: whether the only child in the family, different grade, the different death experience source, different situations of the home to talk about death, presence of internship score has statistical significance (P < 0.05), the only child in the family, freshman, death experience comes from parents, home, never talk, no internship experience of nursing undergraduates hospice care knowledge score higher.

Pain control dimension: the score of religious belief was statistically significant (P < 0.05), and the score of hospice care knowledge of nursing undergraduates without religious belief was higher.

Dimension of social, psychological and spiritual support: The scores of different genders, whether they are the only child, different grades, different sources of death experience, different situations of talking about death in the home and different channels for obtaining knowledge of hospice care were statistically significant (P < 0.05). The scores of male, the only child, the freshman, the death experience from parents, the undergraduate nursing students who never discussed death in the home and obtained knowledge of hospice care from expert lectures were higher.

Therefore, there were statistically significant differences in hospice knowledge scores among nursing undergraduates with different genders, religious beliefs, only child, internship experience, grades, sources of death experience, different situations of talking about death at home, and different channels of obtaining hospice knowledge (P < 0.05). Results are shown in Table 3-4.

**Table 3.**
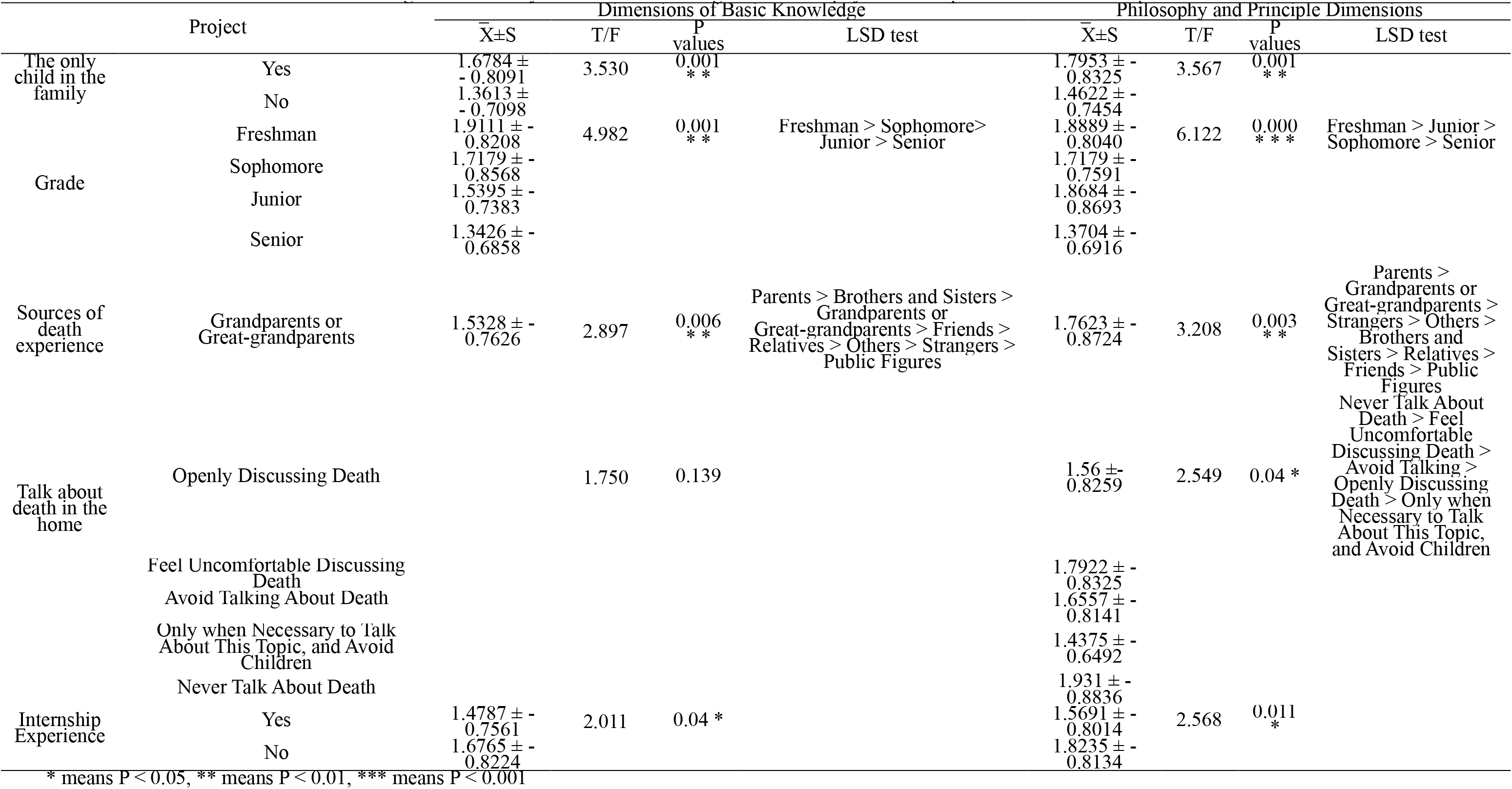
Single Factor Analysis of Basic Knowledge and Philosophy and Principle Dimensions of Hospice Care

**Table 4.**
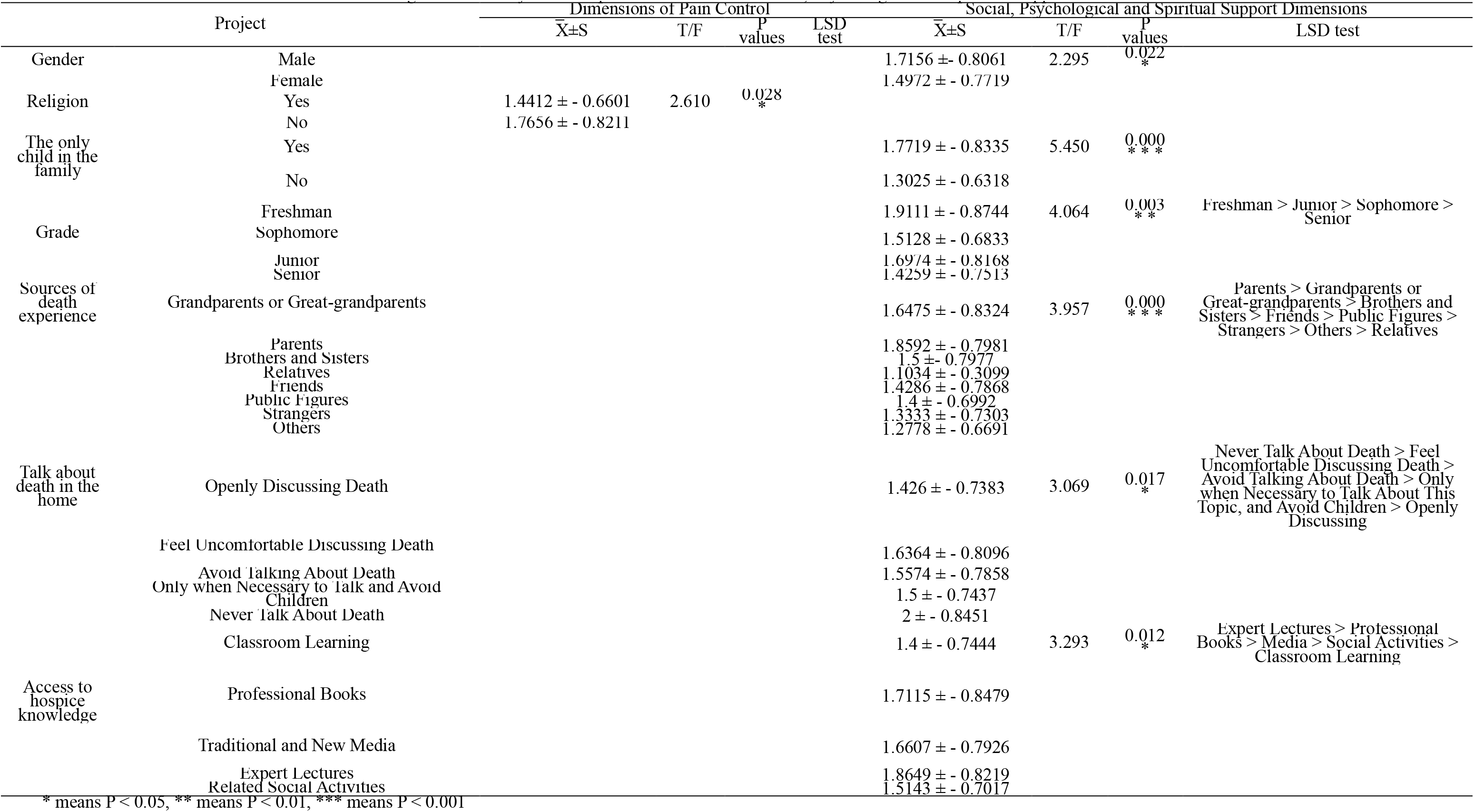
Single Factor Analysis of Hospice Pain Control and Social, Psychological and Spiritual Support Dimensions

#### 2.2.3 Multivariate Linear Regression Analysis of Hospice Care Knowledge Among Undergraduate Nursing Students

Eight variables with statistical significance in the one-way analysis of variance (ANOVA): gender, religion, the only child, internship experience, grade, source of death experience, talk about death in the home, and access to hospice care knowledge were taken as the independent variables, and the score of the hospice care knowledge questionnaire was taken as dependent variable. Multiple linear regression analysis was performed. Results are shown in Table 5. The test level α _in_ =0.05, and α _out_ =0.10.

**Table 5.** Variable Assignments

| Variable | Assignment |
| --- | --- |
| Gender | Male= 0, Female= 1 |
| Grade | Freshman =0, Sophomore =1, Junior =2, Senior =3 |
| Religion | No =0, Yes =1 |
| The only child<br>in the family | No =0, Yes =1 |
| Internship<br>Experience | No =0, Yes =1 |
| Sources of<br>Death<br>Experience | Grandparents or Great-grandparents=0000000, Parents =0000001,<br>Brothers and Sisters=0000010, Relatives =0000100, Friends =0001000,<br>Public Figures =0010000, Strangers =0100000, Others =1000000 |
| Talk About<br>Death in the<br>Home | Openly Discussing Death=0000, Feel Uncomfortable Discussing<br>Death=0001, Avoid Talking About Death=0010, Only when Necessary to<br>Talk and Avoid Children=0100, Never Talk About Death =1000 |
| Access to<br>Hospice<br>Knowledge | Classroom Learning =0000, Professional Books =0001, Media =0010,<br>Expert Lectures =0100, Social Activities =1000 |

The results showed that there were two independent influencing factors of hospice care knowledge among nursing undergraduates: grade and only child. From the perspective of standardized regression coefficient, the influence degree of hospice care knowledge score of nursing undergraduate students from big to small was: freshman > sophomore > junior > senior, the only child > not the only child(Table 6).

**Table 6.**
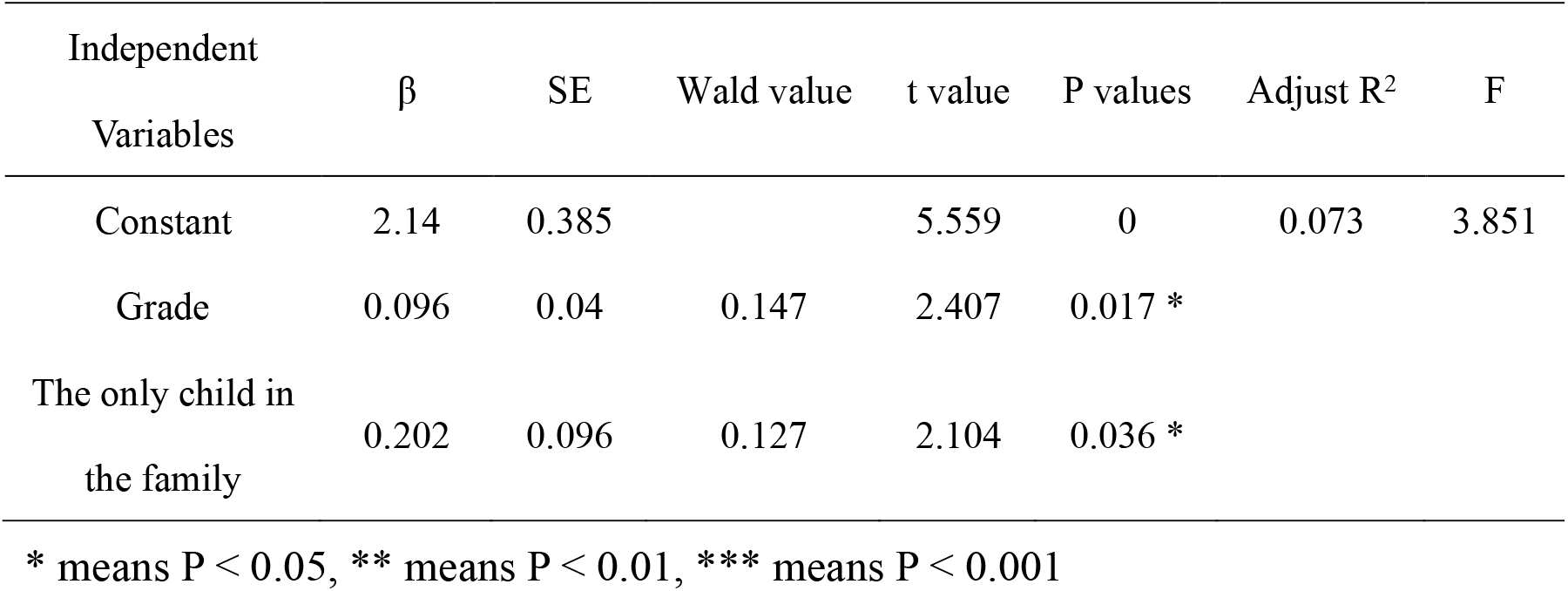
Multiple Linear Regression of Hospice Knowledge Questionnaire for Nursing Undergraduates

### 2.3 Analysis of Hospice Attitude Questionnaire

#### 2.3.1 Score of Hospice Care Attitude Questionnaire for Nursing Undergraduates

There are 19 questions in this questionnaire, including 10 forward questions and 9 reverse questions. Positive questions: 5 points for strongly agreeing, 4 points for agreeing, 3 points for not sure, 2 points for disagreeing, 1 point for strongly disagreeing, and the score for reverse questions is completely opposite, with a total of 95 points. In all valid questionnaires, the highest score was 84, the lowest score was 39, and the mean score was 62.5931±7.2934(Table 7).

**Table 7.** Score of Hospice Attitude Questionnaire

| Dimension | Mean Score | Standard Deviation |
| --- | --- | --- |
| Attitude Evaluation | 27.0379 | 5.6662 |
| Service Object Demand Evaluation | 16.2172 | 2.0522 |
| Personal Death Attitude Evaluation | 19.3379 | 3.1562 |

#### 2.3.2 Single Factor Analysis of Influencing Factors of Hospice Care Attitude of Nursing Undergraduates

The one-way analysis of variance was used to analyze the three dimensions of the hospice care attitude questionnaire: Attitude evaluation, Service object demand evaluation and Personal death attitude evaluation scores.

Dimension of attitude evaluation: The scores of different gender, different age, the only child, different grade, different sources of death experience, talk about different situations of death in the home, internship experience, and different channels to obtain hospice care knowledge were statistically significant (P < 0.05). The scores of female, not the only child, senior, the death experience from strangers, the ability to talk about death in the home openly, the knowledge to obtain hospice care from classroom learning, have the internship experience, and the hospice care attitude of nursing undergraduates aged 18–22 were higher.

Dimension of service object demand evaluation: the scores of whether the patient was the only child, different grades and different sources of death experience were statistically significant (P < 0.05). The scores of hospice care attitude of nursing undergraduates who were not the only child, were in senior year, and the death experience was derived from relatives were higher.Dimension of personal death attitude evaluation: whether the undergraduate was the only child or not and the scores from different death experience sources were statistically significant (P < 0.05); the undergraduate nursing students who were the only child and the death experience sources from their parents had higher scores in hospice care attitude.

Therefore, the scores of hospice care knowledge among undergraduate nursing students who were from different gender, different age, whether they were the only child, whether they had internship experience, different grade, different sources of death experience, different situations of talking about death in their homes and different channels for obtaining hospice care knowledge were statistically significant (P < 0.05). However, there was no significant difference in the knowledge scores of hospice care among nursing undergraduates with different religious beliefs. The results are shown in Table 8–9.

**Table 8.**
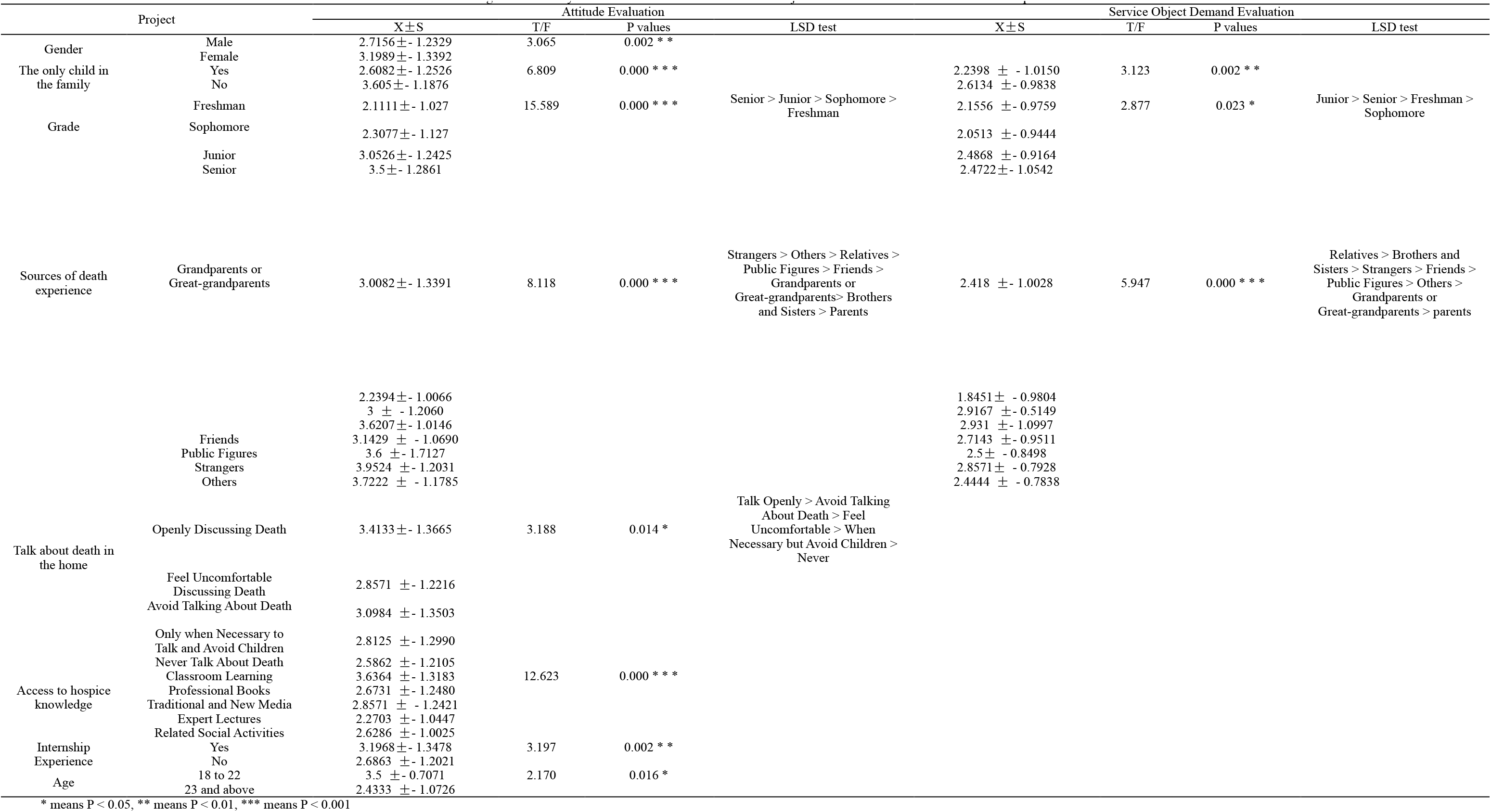
Single Factor Analysis of Attitude Evaluation and Service Object Demand Evaluation Dimensions of Hospice Care

**Table 9.**
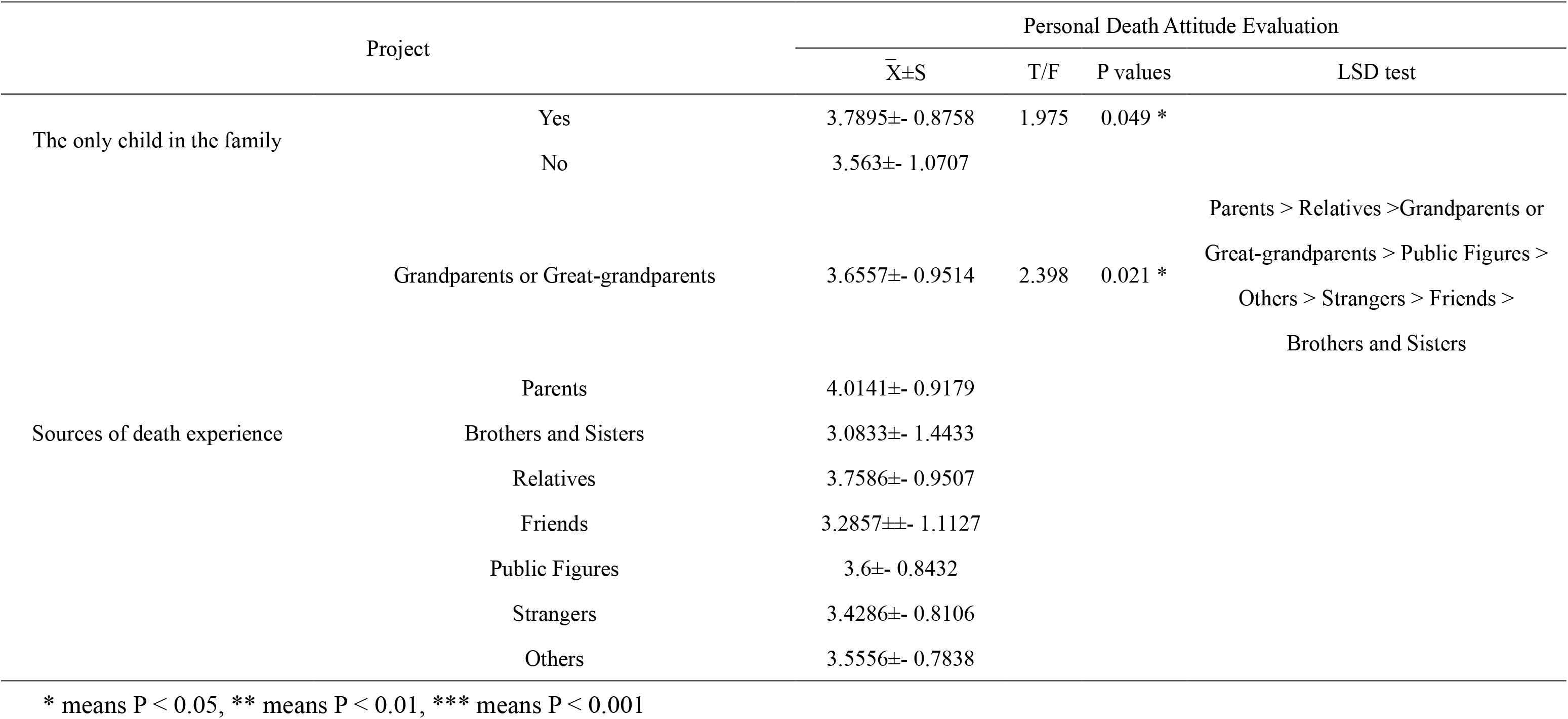
Single Factor Analysis of the Evaluation Dimension of Personal Attitude Towards Death in Hospice Care

#### 2.3.3 Multiple Linear Regression Analysis of Independent Influencing Factors of Hospice Care Attitude of Nursing Undergraduates

Statistically significant in the single factor analysis of variance of variables: gender, age, the only child in the family, internships, grade, death experience source, home to talk about death case, access to hospice care knowledge channel 8 variables as independent variables, such as hospice care attitude questionnaire scores as the dependent variable, using multiple linear regression analysis, variable assignment, see table 10 level of α_in_= 0.05, α_out_ = 0.10.

**Table 10.**
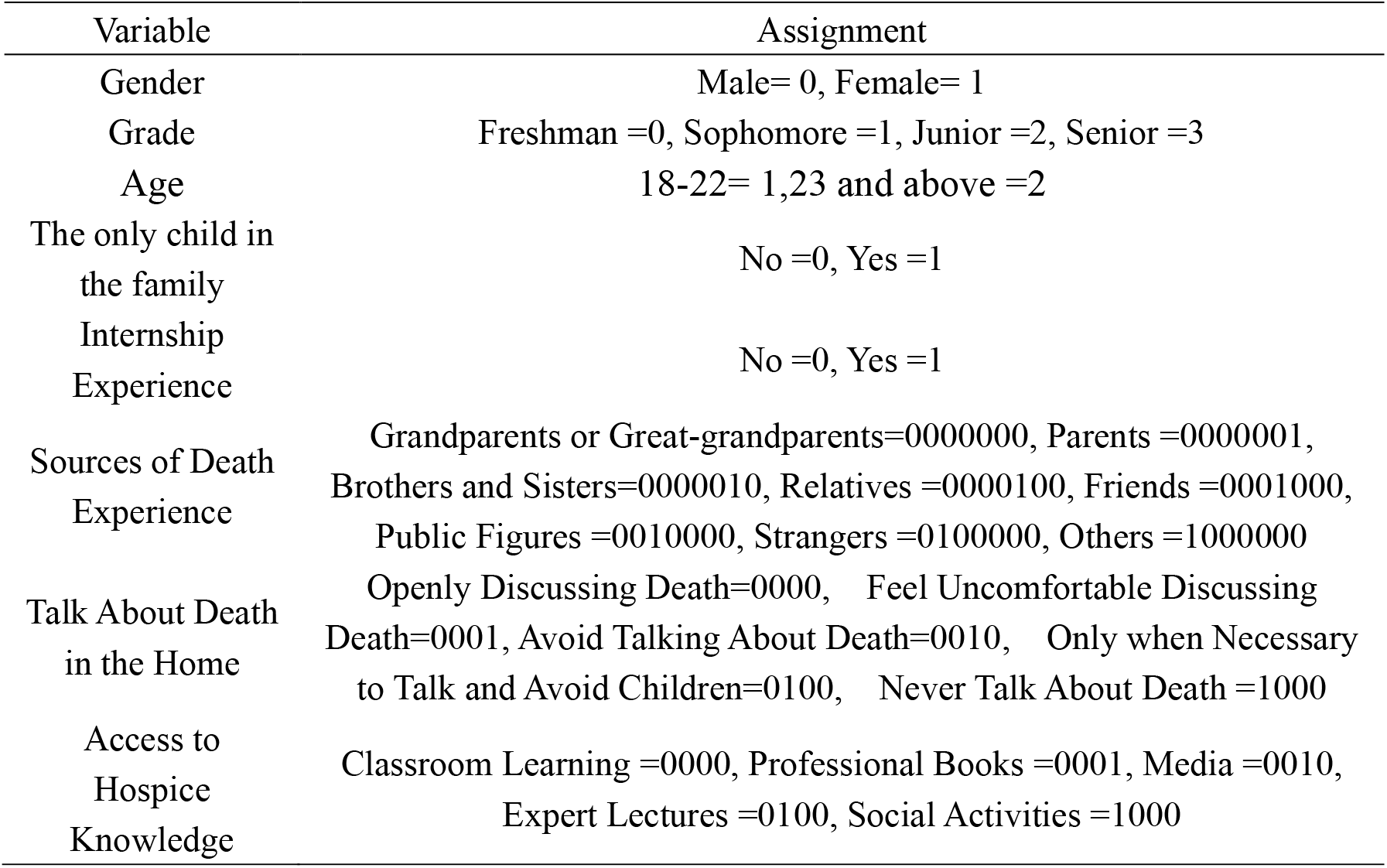
Variable Assignments

| Variable | Assignment |
| --- | --- |
| Gender | Male= 0, Female= 1 |
| Grade | Freshman =0, Sophomore =1, Junior =2, Senior =3 |
| Age | 18-22= 1, 23 and above =2 |
| The only child in the family | No =0, Yes =1 |
| Internship Experience | No =0, Yes =1 |
| Sources of Death Experience | Grandparents or Great-grandparents=0000000, Parents =0000001, Brothers and Sisters=0000010, Relatives =0000100, Friends =0001000, Public Figures =0010000, Strangers =0100000, Others =1000000 |
| Talk About Death in the Home | Openly Discussing Death=0000, Feel Uncomfortable Discussing Death=0001, Avoid Talking About Death=0010, Only when Necessary to Talk and Avoid Children=0100, Never Talk About Death =1000 |
| Access to Hospice Knowledge | Classroom Learning =0000, Professional Books =0001, Media =0010, Expert Lectures =0100, Social Activities =1000 |

The results showed that there were three independent factors influencing the attitude of hospice care in nursing undergraduates: grade, only child and access to hospice care knowledge. From the influence coefficient, the degree of influence of hospice care attitude score of nursing undergraduates is as follows: senior year > junior year > sophomore year > freshman year, not the only child > the only child, classroom learning > professional books > media > expert lectures > social activities(Table 11).

**Table 11.** Multiple Linear Regression of Hospice Attitude Questionnaire for Nursing Undergraduates

| Independent Variables | $\beta$ | SE | Wald value | t value | P values | Adjust R<sup>2</sup> | F |
| --- | --- | --- | --- | --- | --- | --- | --- |
| Constant | 0.403 | 0.884 |  | 0.455 | 0.649 | 0.316 | 16.316 |
| Gender | 0.287 | 0.061 | 0.262 | 4.73 | 0.000 * * * |  |  |
| Age | 0.096 | 0.038 | 0.139 | 2.557 | 0.011 * |  |  |
| The only child in the family | 0.521 | 0.142 | 0.199 | 3.669 | 0.000 * * * |  |  |
| Sources of Death Experience | 0.179 | 0.05 | 0.194 | 3.605 | 0.000 * * * |  |  |

\* means $P < 0.05$ , \*\* means $P < 0.01$ , \*\*\* means $P < 0.001$

## 3. Discussion and Analysis

### 3.1 Lack of Hospice Care Knowledge in Nursing Undergraduates

The results of hospice knowledge questionnaire showed that the highest score of all valid questionnaires was 18, the lowest score was 0, the average score was 9.6793±4.7340, and the average accuracy rate was 53.77%, which was lower than the results of Hou Chen hui’s study^[5]^61.6% and meng Na study results^[45]^Of 60%.The highest correct rate was social support, psychological support and spiritual support (1.7474±0.8114), followed by philosophy and principle (1.7166±0.8116), pain control (1.65861±0.8251). The dimension with the lowest accuracy was basic knowledge dimension (1.6448±0.8218).It shows that nursing undergraduates lack the philosophy and principles of hospice care and the knowledge of hospice care to control pain, and the basic knowledge of hospice care needs to be strengthened.

### 3.2 Nursing Undergraduates Have a Positive Attitude Towards Hospice Care

The highest score was 84, the lowest score was 39, and the mean score was 62.5931±7.2934.Higher than Hou Chen hui^[15]^53.3662±3.4879 and Meng Na^[13]^58.3678 + / - 6.4691.The dimension with the highest accuracy was attitude evaluation (27.0379±5.6662), followed by personal death attitude evaluation (19.3379±3.1562), and the dimension with the lowest accuracy was service object demand evaluation (16.2172±2.0522).The results showed that nursing undergraduates were more positive in attitude evaluation and personal attitude toward death evaluation.

### 3.3 Influencing Factors of Hospice Cognition Among Nursing Undergraduates

#### 3.3.1 Different Grades of Nursing Undergraduates Have Different Cognition of Hospice Care

The results of multi-factor analysis of hospice knowledge questionnaire for nursing undergraduates showed that the scores of hospice knowledge of nursing undergraduates in different grades were statistically significant (P =0.017).The results of multi-factor analysis of hospice attitude questionnaire for nursing undergraduates showed that the scores of hospice attitude of nursing undergraduates in different grades were statistically significant (P =0.000).The results of single factor analysis showed that first-year nursing undergraduates scored higher than other grades in basic knowledge dimension, philosophy and principle dimension, social support, psychological support and spiritual support dimension (Table 3-4), which was different from previous studies^[14-16]^May be related to the different time when schools offer hospice care courses. And freshman nursing undergraduates’ attitude toward hospice care attitude evaluation and service object demand evaluation dimension scores are for the bottom (Table 3,4), freshman nursing undergraduates hospice negative attitude may contact with not true hospice care clinical environment, and influenced by traditional thought, of the unknown true lack of perceptual knowledge about the status of the hospice care Therefore, the school advocates scenario simulation teaching, which can reduce students’ fear of the unknown to the real clinical situation, increase their perceptual knowledge and improve their attitude to practice by simulating the real environment. Senior grade nursing undergraduate students basic knowledge of hospice care knowledge dimension, philosophy and principles of dimension and the control dimension scores were the lowest pain, social support, psychological support and spirit support dimension score the penultimate (Table 8-9), with most of the senior grade nursing undergraduate students all want to go to the hospital clinical practice, training hospital for more general hospital, less contact with hospice care, Therefore, during the internship, both individuals and schools should pay attention to the review and application of what they have learned in the past, especially to review non-clinical common knowledge.

#### 3.3.2 Whether the Only Child Affects the Hospice Cognition of Nursing Undergraduates

From the results of multiple linear regression analysis of hospice knowledge questionnaire of nursing undergraduates, whether the only child of nursing undergraduates had statistical significance on hospice knowledge score (P=0.036);The results of multiple linear regression analysis of hospice care attitude questionnaire for nursing undergraduates showed that whether the only child of nursing undergraduates had statistical significance on hospice care attitude score (P=0.000).The results of univariate analysis show that the score of hospice care knowledge of only children in all dimensions is higher than that of non-only children (see Table 3-4), which may indicate that families with only children are under greater pressure to provide for the aged^[17]^To be more sensitive to hospice care knowledge;The score of the evaluation dimension of the attitude towards death of the only child in hospice care is higher than that of the non-only child, indicating that the only child has a more positive attitude towards death^[18]^, may be related to the growth environment of the only child and the non-only child. However, the attitude evaluation and service object demand evaluation dimensions of non-only children in hospice care are more positive (see Table 8-9), which is similar to meng Na’s^[14]^The research results are consistent, and may be with non-only child family pension pressure is less^[17]^And lower levels of death anxiety.

#### 3.3.3 Nursing Undergraduates with Different Access to Hospice Knowledge Had Different Attitudes Towards Hospice Care

The results of multiple linear regression analysis of hospice attitude questionnaire for nursing undergraduates showed that the hospice attitude of nursing undergraduates with different access to hospice knowledge was statistically significant (P =0.000), according to the results of single factor analysis on classroom learning for hospice care knowledge of nursing undergraduates in hospice care attitude questionnaire of evaluation and evaluation dimension scores the highest service object demand, shows the two dimension of hospice care attitude more positive than other channels, might be more full and accurate, and the classroom learning is beneficial to cultivate nursing undergraduates correct view on death; And get hospice care knowledge from experts lecture nursing undergraduates personal attitude evaluation dimension scores the highest death, may be due to the expert lectures more into the instance and a targeted on one hand, in hospice care and involve the issue of life and death to life, easy to attract the audience attention and resonate, heard expert lectures of nursing undergraduates so set up the positive attitude towards death.

## 4. Conclusion

Among the medical staff, nursing staff contact with terminal patients for longer time, is the main force of the nursing staff, literature shows that the degree of hospice care cognition of nursing staff directly affects the quality of hospice care service^[19-25]^.And undergraduate nursing students are engaged in hospice nursing with higher education, their hospice cognition should not be ignored.

This survey found that nursing undergraduates lack of hospice care knowledge, lack of hospice care philosophy and principles and hospice care pain control knowledge, the grasp of hospice basic knowledge needs to be strengthened. The attitude of nursing undergraduates towards hospice care was more positive, especially in the evaluation of attitude and personal attitude towards death. The influencing factors of hospice care knowledge for nursing undergraduates include gender, religious belief, only child, practice experience, grade, source of death experience, the situation of talking about death at home, and channel of obtaining hospice care knowledge, among which grade and only child are independent influencing factors. The influencing factors of hospice care attitude of nursing undergraduates included gender, age, only child, internship experience, grade, source of death experience, the situation of talking about death at home, and access to hospice care knowledge channel, among which grade, only child and access to hospice care knowledge channel were independent factors.

In order to enhance the hospice awareness of nursing undergraduates and cultivate more highly educated and high-quality talents for hospice cause, on the one hand, hospice education should be strengthened, and hospice courses should be reasonably set up both theoretically and practically in the teaching plan of nursing undergraduates^[26]^To improve the teaching methods and assessment system, enrich the classroom content, in order to mobilize students’ learning enthusiasm, strengthen the nursing undergraduates’ knowledge of hospice care, establish a correct view of death, so as to improve their hospice care cognition level. On the other hand, we should strive for the support of the government, and strive for the support of the government policy is conducive to the promotion of hospice care service, which is conducive to improving the cognitive level of hospice care in the whole society, promoting the development of hospice care education, and improving the cognitive level of hospice care in nursing undergraduates.

## Data Availability

All data produced in the present study are available upon reasonable request to the authors
All data produced in the present work are contained in the manuscript

http://www.cnaflc.org/lzgh/14291.jhtml

## Notes

### Competing Interest Statement

The authors have declared no competing interest.

### Funding Statement

This study did not receive any funding

### Author Declarations

Ethical approval was granted by ethics Committee of Beijing University of Chinese Medicine

